# Social isolation, loneliness and all-cause dementia: a longitudinal and imaging-genetic study in the UK Biobank cohort

**DOI:** 10.1101/2021.06.30.21259818

**Authors:** Chun Shen, Barbara J. Sahakian, Wei Cheng, Jujiao Kang, Guiying Dong, Chao Xie, Xing-Ming Zhao, Jianfeng Feng

## Abstract

**INTRODUCTION:** Current findings of the relative influence of social isolation and loneliness on dementia are contradictory, and the potential neurobiological mechanisms are unclear.

**METHODS:** We utilized the UK Biobank to investigate the relationships of social isolation and loneliness with dementia (n = 462,619). Neuroanatomical correlates were identified in a subset of participants (n = 32,263). The transcriptomic signatures of related brain changes were characterized by gene enrichment analysis.

**RESULTS:** After full adjustment, social isolation but not loneliness was associated with dementia (hazard ratio: 1.28, 95% confidence interval: 1.17-1.39). Isolated individuals had reduced gray matter volumes in temporal, frontal, occipital and subcortical regions (e.g., hippocampus and amygdala). Relevant brain changes were spatially correlated with genes involved in mitochondrial dysfunction and oxidative phosphorylation, and down-regulated Alzheimer’s disease-related genes.

**DISCUSSION:** Social isolation is an independent risk factor for dementia, which could be partly explained by related structural changes coupling with altered molecular functions.

## 1. BACKGROUND

Social isolation (objective reflection of social relationships) and loneliness (perceived social isolation) are serious yet underappreciated public health problems that are particularly associated with old age.^1^ Dementia is a major cause of disability in the elderly, affecting over 46 million people worldwide in 2015 and was estimated to increase to 131.5 million by 2050.^2^ To date, the influence of social isolation and loneliness on dementia is still unclear. Whereas in some studies social isolation but not loneliness was associated with increased risk of dementia and cognitive decline,^3, 4^ other studies found the opposite result.^5^ One possibility for this discrepancy is that the associations may be impacted by risk factors which were not consistently considered in past studies. To what extent established risk factors account for such associations requires further elucidation.

Moreover, little is known about the underlying neurobiological mechanisms. The social brain hypothesis posits that the evolution of human brain is driven by increasingly complex social selection pressures.^6^ Thus, social isolation may impair the brain regions involved in higher cognitive and affective processes, and in turn affect cognitive abilities. Only a paucity of studies have explored the neural underpinnings of social isolation and loneliness. Structural and functional changes in several brain regions including prefrontal, temporal and parietal cortices, amygdala, hippocampus, striatum and ventral tegmental area were reported.^7-12^ Spreng et al. (2020)^13^ reported that loneliness is associated with the default network. However, findings are mixed probably due to the heterogeneity of methods and study design.^14^

Social factors may also play a significant role in regulating the transcriptional activity of human genome.^15^ Previous gene expression studies in post-mortem nucleus accumbens and dorsolateral prefrontal cortex revealed that loneliness-related differentially expressed genes were associated with Alzheimer’s disease (AD) and immune dysfunction.^16, 17^ One recent approach leveraged brain-wide gene expression atlases to link molecular function to macroscale brain organization,^18^ which was helpful to understand disease-related brain alterations and has been used in depression^19^ and neurodegenerative diseases.^20^

The present study utilized the UK Biobank cohort to examine the relative influence of social isolation and loneliness on incident dementia, and to quantity the extent to which these associations were explained by various risk factors such as biological and psychological factors. Next, we investigated the neuroanatomical correlates of social isolation and loneliness, and tested whether the identified brain alterations could mediate the prospective associations of social isolation and loneliness with cognitive abilities. Finally, using the Allen Human Brain Atlas (AHBA) microarray dataset,^21^ we analyzed the gene expressions of which biological processes or functional pathways were associated with social isolation- and loneliness-related brain changes. Besides, we specifically examined the associations between brain changes and AD-related genes.

## 2. METHODS

### 2.1. Participants

The UK Biobank is a prospective epidemiological study that involves over 500,000 individuals recruited in 22 centers across the UK between 2006 and 2010.^22^ The study has collected extensive questionnaire data, physical measurements, and biological samples. A subset of the cohort has been invited back to collect multimodal imaging data and repeat behavioral assessments since 2014. All participants are followed up for health conditions through linkage to national electronic health-related datasets. The health follow-up data used in the present study started at enrollment and ended in January 2021. All participants provided informed consents and the ethical approval was from the North West Multi-Centre Research Ethics Committee.

### 2.2. Defining social isolation and loneliness

Social isolation and loneliness were calculated from scales that were used in previous studies.^23^ Social isolation was assessed with three questions: number of people living together in the household (1 = living alone), frequency of visiting friends or family or having they visit you (1 = less than once a month) and attending leisure or social activities at least weekly (1 = no participation). An individual was defined as socially isolated if he or she scored 2 or 3, and those who scored 0 or 1 were classified as not isolated. Loneliness was constructed from two questions: often feeling lonely (1 = yes) and frequency of confiding in close people (1 = less than once every few months). An individual was defined as lonely if he or she scored 2, and not lonely if he or she scored 0 or 1.

### 2.3. Ascertainment of dementia

All-cause dementia was identified as International Classification of Diseases 10th codes F00, F01, F02, F03 or G30 of the “first occurrence” data-fields generated by the UK Biobank (data version: January 2021; https://biobank.ndph.ox.ac.uk/showcase/label.cgi?id=1712), which were ascertained by the combination of primary care, hospital in-patient, death register and self-reported data. Date of diagnosis was set as the earliest date of dementia codes recorded regardless of source used. Prevalent dementia cases were defined as the date of diagnosis occurred within the first 3 years of follow-up or self-reported “dementia, AD or cognitive impairment” at baseline, and were excluded to avoid the possible reverse-causation bias (n = 494). The source of incident all-cause dementia cases was reported in Table S1.

### 2.4. Assessment of cognitive performance

Pairs matching and reaction time tests were administered to almost all participants at baseline and over 40,000 participants at the imaging visit. In the first test, respondents were asked to correctly identify matches from six pairs of cards after they had memorized their positions. The number of incorrect matches was recorded, with a greater number reflective of a poorer visual memory. This visual memory error score was logarithmically transformed (In(x+1)) due to skewed distribution and zero-inflation. The card-game “Snap” asked participants to press a button when two simultaneously presented cards matched, with shorter reaction time represented faster speed of processing. The log-transformed mean reaction time over 12 rounds was used (In(x)). In addition, these two tests were normalized and then averaged to yield a composite score of cognition.

### 2.5. Structural MRI data

Neuroimaging data were collected across three dedicated, identical imaging centers. The detailed MRI acquisition protocol has been described elsewhere.^24, 25^ All structural MRI data were preprocessed in the Statistical Parametric Mapping software version 12 using the CAT12 toolbox with default settings, including the usage of high-dimensional spatial normalization with an already integrated Dartel template in Montreal Neurological Institute (MNI) space. All images were subjected to nonlinear modulations and corrected for each individual head size. Images were then smoothed with an 8mm full-width at half-maximum Gaussian kernel with the resulting voxel size of 1.5mm^3^. We focused our analyses within the automated anatomical labelling atlas 3^26^ excluding cerebellum.

### 2.6. Transcriptomic data

We used the transcriptomic data from six neurotypical adult brains in the AHBA.^21^ Tissue samples in left hemisphere were used due to the right hemisphere data were only available for two donors. The preprocessing steps included probe-to-gene re-annotation, intensity-based data filtering, probe selection by mean, separating tissue samples into subcortical and cortical regions, and within-donor normalization, as reported previously.^27^ Finally, we obtained expression values for 15,408 unique genes at 711 cortical and 135 subcortical locations separately.

### 2.7. Statistical analysis

#### 2.7.1. Cox proportional hazard model

Associations of social isolation and loneliness with dementia incidence were investigated using Cox proportional hazard models with age as a timescale. The results were presented as hazard ratios (HRs) and 95% confidence intervals (CI). The proportional hazard assumptions were checked using Schoenfeld residuals, and no major violations were observed. We restricted analyses to participants who had complete data on baseline social isolation, loneliness and cognitive tests, and incident dementia. The minimal model was adjusted for age, sex and ethnicity. To assess the extent to which other risk factors explained the associations, percentage of excess risk mediated (PERM) was calculated for: (1) socioeconomic factors (education and household income); (2) biological factors (BMI, blood pressure, chronic illness and *APOE* genotype); (3) cognitive factors; (4) health behavior (alcohol intake, current smoker and physical activity); (5) depressive symptoms; (6) loneliness (for social isolation as the exposure) or social isolation (for loneliness as the exposure). Covariates were treated as categorical variables with missing value as a separate category (Method S1 and Table S2). Finally, a full model including all risk factors was conducted. PERM was estimated using the formula: [HR_(age, sex and ethnicity adjusted)_ – HR_(age, sex, ethnicity and risk factor adjusted)_]/[HR(age, sex and ethnicity adjusted) –1] ⍰ 100. ^28^ We also performed subgroup analyses to assess potential modification effects by sex (men or women) and age (< 60 or ⍰ 60 years), and a sensitivity analysis using complete cases to test the robustness. Calculations were performed by R version 3.6.0 using the survival package.^29^

#### 2.7.2. Whole-brain and voxel-wise analyses

Linear regression models were conducted to investigate the cross-sectional associations of social isolation and loneliness with gray matter volume (GMV) separately. Age, education and household income at the imaging visit, and sex, ethnicity, site and total intracranial volume (TIV) were used as covariates of no interest. Multiple comparison correction was performed at voxel level with a *P*_FDR_ < .05. A significant cluster was defined on the basis of an 18-connectivity criterion^30^ and having more than 217 voxels falling into the 90% CI of the smoothing kernel voxels.^27^ Using the data-driven approach, we found significant GMVs associated with social isolation but no regions for loneliness. Three sensitivity analyses were performed: (1) excluding participants with dementia at any time (n = 29), (2) additionally adjusted for loneliness, (3) additionally adjusted for depressive symptoms. The identified brain regions were decoded for cognitive functions using Neurosynth meta-analysis toolbox.^31^ We calculated cross-voxel Spearman’s correlations between the significant social isolation *t*-statistic map and the association *Z*-statistic maps. The top 100 negative terms were selected, from which the anatomical terms such as “hippocampus” were removed, and the retained cognitive terms were visualized on a word-cloud plot (Method S2). Additionally, we tested the cross-sectional relationship between the significant GMVs and cognitive performance in the UK Biobank controlling for the same covariates as in the whole-brain analysis. Finally, the mediation effect of the significant GMVs on the association between social isolation at baseline and cognitive performance at the imaging visit was examined. Age, education and household income at baseline, time interval between baseline assessment and imaging collection, and sex, ethnicity, site and TIV were adjusted for. The significance of the mediation was estimated by 10,000 bias-corrected bootstrapping, which was performed using the mediation toolbox developed by Wager et al..^32^

#### 2.7.3. Transcriptomic analysis

Partial least square (PLS) regression was used to relate the social isolation *t*-statistic map (response variable) to the gene expression data (predictor variable). The response variable was calculated by the average *t*-value of a spherical region with a radius of 4mm centered by the MNI coordinates of each gene expression sampling site.^27^ PLS was performed for cortical and subcortical regions separately. The first PLS component (PLS1) was the linear combination of gene expression values maximizing the covariance between the expression profile and GMV changes. The statistical significance of the variance explained by PLS1 was tested by recomputing PLS using 5000 null *t*-statistic maps which were obtained by label shuffling for social isolation, and counting the number of times the explained variance was higher than the original observation (denoted as *P*_perm_).^33^ As PLS1 was not significant in subcortical regions, the following analyses were limited in cortical regions.

Bootstrapping (5000 times) was used to estimate the variability of each gene’s PLS1 weight, and the ratio of the weight to its bootstrapped standard error was used to calculate the *Z*-score.^34^ Genes with PLS1 weights *Z* > 4 (PLS1+) or *Z* < -4 (PLS1-) (all *P*_FDR_ < .001) were used to calculate enrichments in both Kyoto Encyclopedia of Genes and Genomes (KEGG) pathways and Gene Ontology (GO) terms of biological processes (Method S3). FDR correction was performed for KEGG pathways and GO biological processes simultaneously. Significance was set at *P*_FDR_ < .05. Specially, we examined whether AD-related dysregulated genes were expressed most in regions that were morphometrically correlated to social isolation. A recent systematic integrated analysis reported 2444 up-regulated and 2978 down-regulated AD-related genes (Method S4).^35^ Furthermore, we investigated the relationship of social isolation-related GMV changes with the average expression level of up- and down-regulated AD-related genes separately. The significance was tested by 5000 times permutation, in which the null distribution was defined by 5000 random *t*-statistic maps described above.

## 3. RESULTS

### 3.1. Demographics

At baseline, 462,619 participants provided complete data on social isolation, loneliness, cognitive tests and incident all-cause dementia (Table 1 and Table S3). Of these, 41,886 (9%) individuals reported as socially isolated, and 29,036 (6%) individuals felt lonely. Compared with controls, both isolated (F(1,462608) = 205.08, *P* < .001) and lonely individuals (F(1,462608) = 41.04, *P* < .001) had lower cognitive ability after controlling for age, sex, ethnicity, education and income. During a mean follow-up of 11.69 years (SD = 1.65), 4998 participants developed dementia. After an average of 8.81 years (SD = 1.72), 32,263 individuals were invited back to participate in the MRI collection, and social isolation, loneliness and demographic information were simultaneously measured (Table S4). At the imaging visit, 2371 (7%) participants were socially isolated and 1503 (5%) participants were lonely.

**Table 1:**
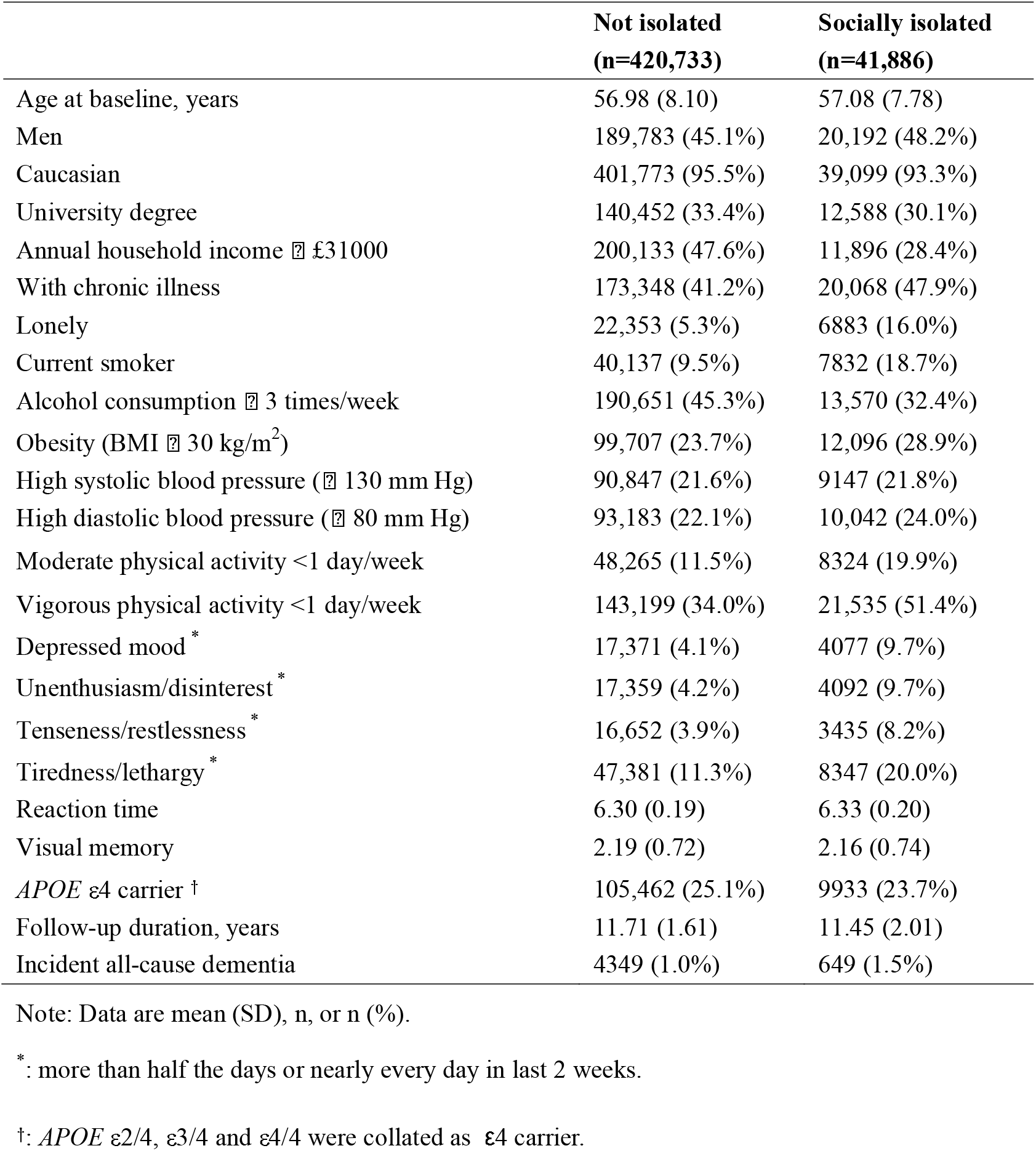
Baseline characteristics of study samples in the UK Biobank cohort (n=462,619)

|  | **Not isolated**<br>**(n=420,733)** | **Socially isolated**<br>**(n=41,886)** |
| --- | --- | --- |
| Age at baseline, years | 56.98 (8.10) | 57.08 (7.78) |
| Men | 189,783 (45.1%) | 20,192 (48.2%) |
| Caucasian | 401,773 (95.5%) | 39,099 (93.3%) |
| University degree | 140,452 (33.4%) | 12,588 (30.1%) |
| Annual household income ≥ £31000 | 200,133 (47.6%) | 11,896 (28.4%) |
| With chronic illness | 173,348 (41.2%) | 20,068 (47.9%) |
| Lonely | 22,353 (5.3%) | 6883 (16.0%) |
| Current smoker | 40,137 (9.5%) | 7832 (18.7%) |
| Alcohol consumption ≥ 3 times/week | 190,651 (45.3%) | 13,570 (32.4%) |
| Obesity (BMI ≥ 30 kg/m<sup>2</sup>) | 99,707 (23.7%) | 12,096 (28.9%) |
| High systolic blood pressure (≥ 130 mm Hg) | 90,847 (21.6%) | 9147 (21.8%) |
| High diastolic blood pressure (≥ 80 mm Hg) | 93,183 (22.1%) | 10,042 (24.0%) |
| Moderate physical activity <1 day/week | 48,265 (11.5%) | 8324 (19.9%) |
| Vigorous physical activity <1 day/week | 143,199 (34.0%) | 21,535 (51.4%) |
| Depressed mood * | 17,371 (4.1%) | 4077 (9.7%) |
| Unenthusiasm/disinterest * | 17,359 (4.2%) | 4092 (9.7%) |
| Tenseness/restlessness * | 16,652 (3.9%) | 3435 (8.2%) |
| Tiredness/lethargy * | 47,381 (11.3%) | 8347 (20.0%) |
| Reaction time | 6.30 (0.19) | 6.33 (0.20) |
| Visual memory | 2.19 (0.72) | 2.16 (0.74) |
| APOE ε4 carrier † | 105,462 (25.1%) | 9933 (23.7%) |
| Follow-up duration, years | 11.71 (1.61) | 11.45 (2.01) |
| Incident all-cause dementia | 4349 (1.0%) | 649 (1.5%) |

Note: Data are mean (SD), n, or n (%).
\*: more than half the days or nearly every day in last 2 weeks.
†: APOE ε2/4, ε3/4 and ε4/4 were collated as ε4 carrier.

### 3.2. Social isolation but not loneliness was associated with incident all-cause dementia

After adjustment for age, sex and ethnicity, the HR for incident all-cause dementia was 1.64 (95% CI: 1.51-1.78) for social isolation compared with no social isolation. Further adjustment for different risk factors attenuated the association (PERM: 8%-31%). The overall attenuation after adjustment for all risk factors was 56% to 1.28 (95% CI: 1.17-1.39; Figure 1). In contrast, the minimally adjusted HR for loneliness was 1.55 (95% CI: 1.40-1.71), but reduced to nonsignificance when all risk factors were included (*P* = .45). Adjustment for depressive symptoms had the greatest impact on the association, which was decreased by 75% (Figure 1).

**Figure 1.**
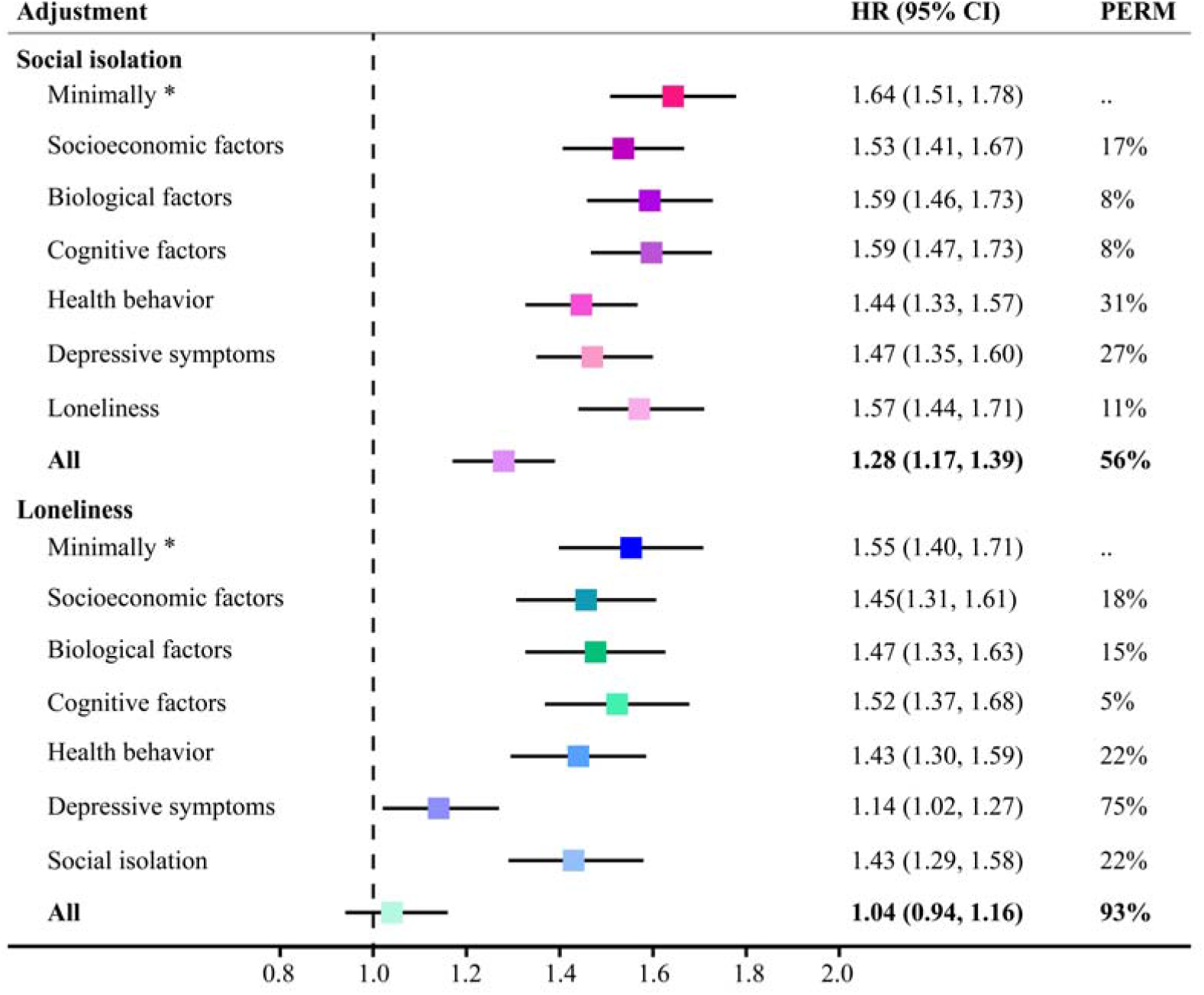
Associations of social isolation and loneliness with incident all-cause dementia, and proportions attributable to different risk factors (n=462,619). HR=hazard ratio. PERM=percentage of excess risk mediated. ^*^: adjusted for age, sex and ethnicity.

Subgroup analyses found the association between social isolation and dementia was consistent across sex (Figure S1). However, the association was only significant in the elderly group with full adjustment (HR: 1.30, 95% CI: 1.18-1.42; Figure S2). Consistent findings were found in the complete case analyses (Figure S3).

### 3.3. Social isolation-related brain structures and the association with cognition

The whole-brain analysis revealed that brain structures encompassing temporal, frontal, occipital and subcortical regions were associated with social isolation (Figure 2A and Table S5). The most significant regions were located in bilateral hippocampus and parahippocampus, left thalamus, fusiform and amygdala, and right supramarginal gyrus. The findings were robust if excluding patients with dementia (Figure S4), and similar brain regions were found if further adjusted for loneliness (Figure S5) or depressive symptoms (Figure S6).

**Figure 2.**
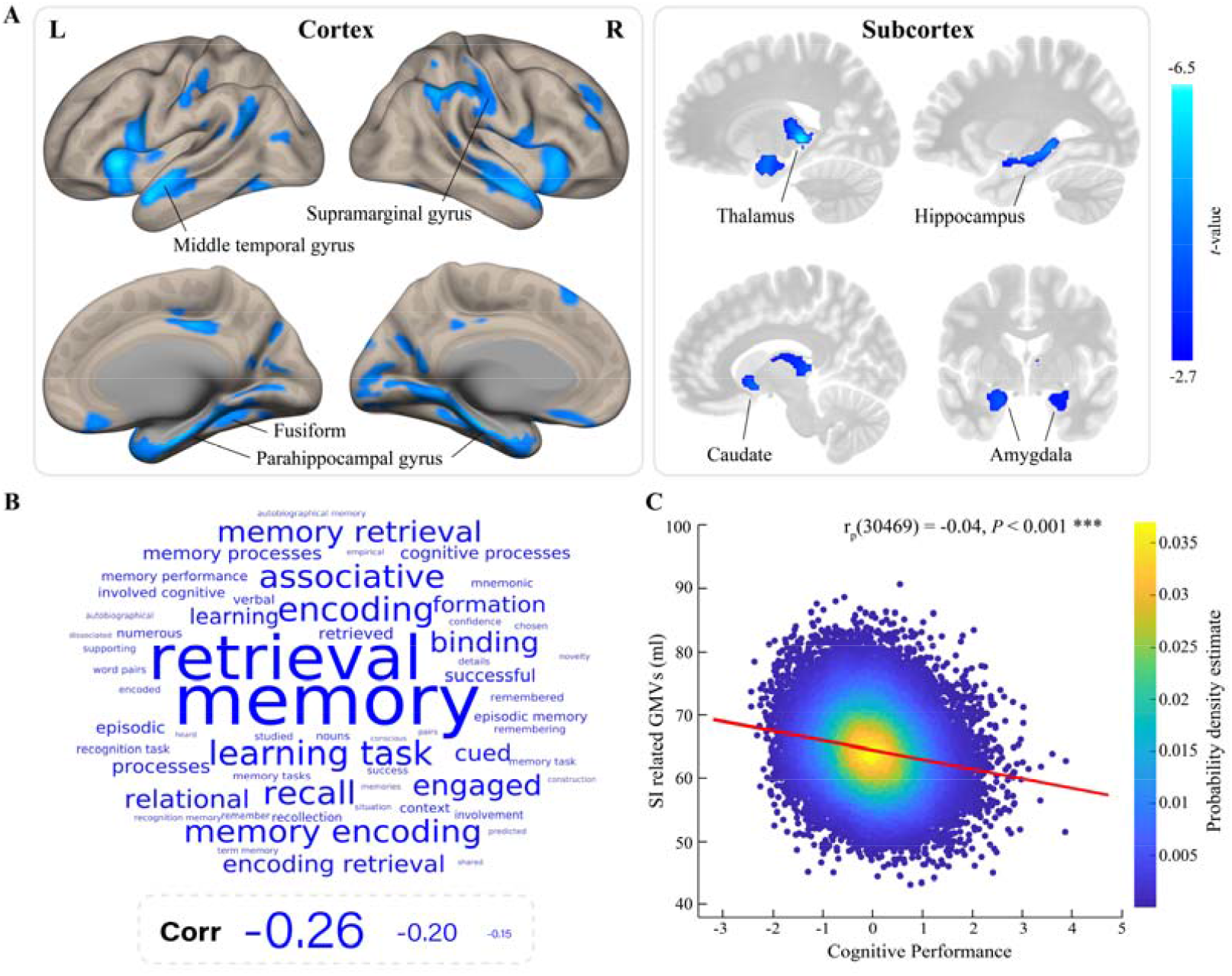
Neuroanatomical correlates of social isolation at the imaging visit (n=32,263). A. Results of whole-brain voxel-wise analysis of social isolation. Age, education and household income at the imaging visit, and sex, ethnicity, site and TIV were used as covariates. Multiple comparison correction was performed at voxel level with a *P*_FDR_ < .05, and a cluster was significant with more than 217 voxels. Significant GMVs in cortical and subcortical regions are presented separately. The color bar presents *t*-value. B. Word-cloud plot of cognitive terms associated with social isolation related GMVs. The size of cognitive terms corresponds to the Spearmen’s correlation of corresponding meta-analytic maps in Neurosynth with the significant social isolation *t*-statistic map. C. Cross-sectional association between social isolation related GMVs and cognitive performance in the UK Biobank (n=30,480). Cognitive performance was a composite score of visual memory and reaction time. The partial correlation coefficient with the same covariates as in the whole-brain analysis is showed. The color bar presents the estimated probability density.

Using Neurosynth, we found the significant regions where GMV was smaller in socially isolated individuals than controls tend to be involved in memory and learning tasks (Figure 2B and Table S6). In the UK Biobank, the total GMV of the significant brain regions was associated with cognitive performance at the imaging visit (r_p_(30469) = -0.04, *P* < .001; Figure 2C), and could significantly mediate the prospective relationship between social isolation and cognitive performance (path ab: β = 0.003, 4% of the total effect size, *P* < .001; Figure 3).

**Figure 3.**
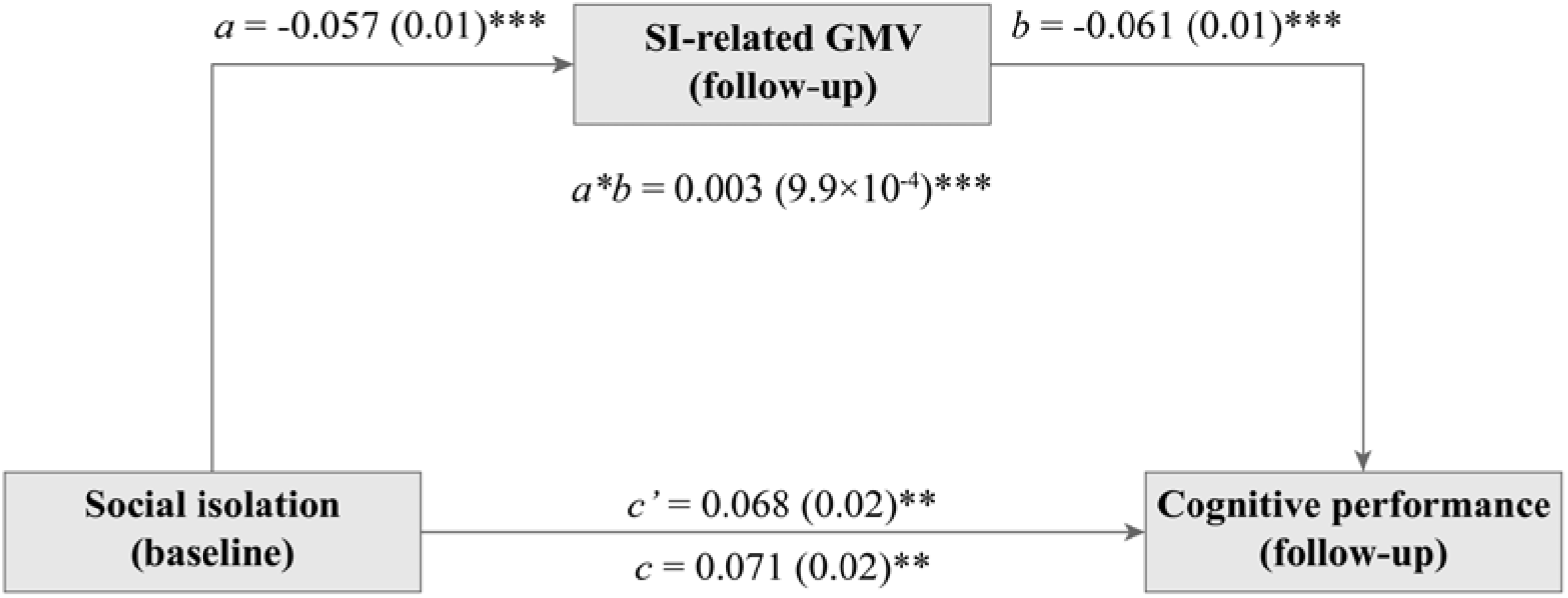
Mediation analysis using the identified gray matter volume as a mediator of the prospective relationship between social isolation and cognitive performance (n=30,612). The predictor in the model is social isolation at baseline, the mediator is the total of significant GMVs associated with social isolation at the imaging visit, and the dependent variable is cognitive performance at the imaging visit, which was a composite score of visual memory and reaction time. Covariates included age, education and household income at baseline, time interval between the baseline assessment and imaging collection, and sex, ethnicity, site and TIV. Path *a* measures the association between the predictor and the mediator; path *b* represents the effect of the mediator on the dependent variable while controlling for the predictor; path *c* measures the total relationship between the predictor and the dependent variable; path *c’* measures the direct effect; the mediation effect is the product of path *a* and path *b* (*a*^*\**^*b*). SI = social isolation. ^**^*P* < .01, ^***^*P* < .001.

### 3.4. Relationship between structural changes and brain gene expression profiles

In cortical regions, the PLS1 was significant (i.e., explained 15% of the variance of social isolation-related structural changes, *P*_perm_ < .05; Result S1). Based on the normalized PLS1 weights, there were 2029 genes in the PLS1+ gene set (*Z* > 4) and 1048 genes in the PLS1-gene set (*Z* < -4) (all *P*_FDR_ < .001; Figure S7 and Table S7). After the FDR correction, the PLS1+ gene set was enriched in 38 biological processes such as “mitochondrion organization” and “oxidative phosphorylation”, and 6 KEGG pathways such as “Parkinson’s disease” and “Alzheimer’s disease” (Figure 4A and Table S8). No significant enrichment results for the PLS1-gene set.

**Figure 4.**
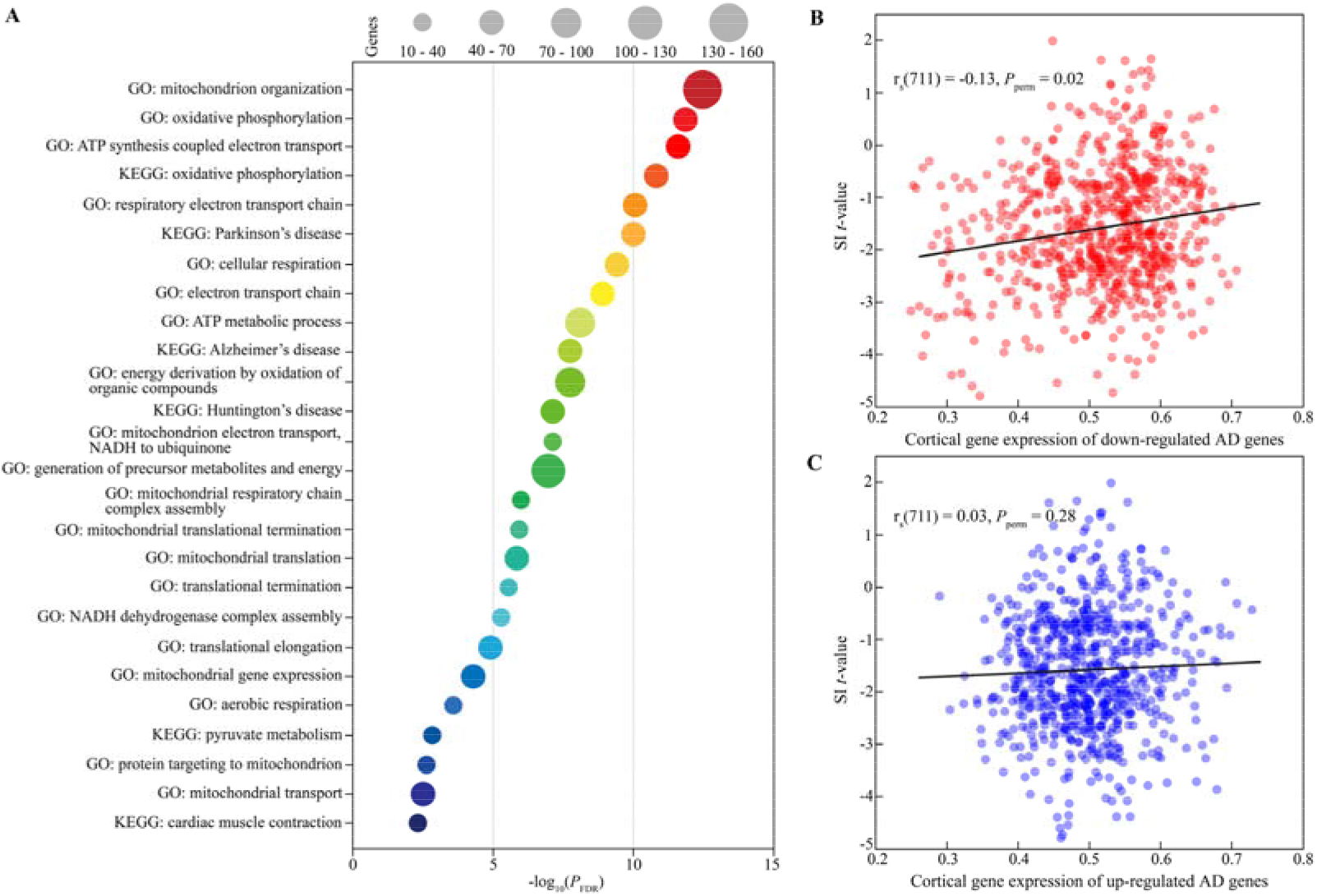
Relationship between social isolation related brain structural changes and gene expression. A. Functional enrichment results of PLS1-genes in cortical regions. GO biological processes and KEGG pathways with a *P*_FDR_ < .005 are presented. The size of the circle represents the number of genes involved in a given ontology term. B. Association between social isolation *t*-value and the average expression level of down-regulated AD-related genes in cortical regions. C. Association between social isolation *t*-value and the average expression level of up-regulated AD-related genes in cortical regions. The significance was tested by 5000 times permutation, in which the null distribution was defined by 5000 random *t*-statistic maps obtained by label shuffling for social isolation.

Moreover, PLS1+ gene set was highly enriched among genes that were recently reported as underexpressed in post mortem brain tissue from patients with AD (*P*_FDR_ < .001; Table S9). Social isolation-related changes in regional GMV was positively associated with cortical gene expression of down-regulated AD-related genes (r_s_(711) = 0.13, *P*_perm_ = .02; Figure 4B). Nevertheless, no significant spatial correlation with up-regulated AD-related genes was found (*P*_perm_ = .28; Figure 4C).

## 4. DISCUSSION

This study suggested that social isolation but not loneliness was associated with elevated risk of incident all-cause dementia. Socially isolated individuals presented reduced GMVs of brain regions involved in memory and learning, which partly mediated the prospective relationship between social isolation and cognitive performance. Transcriptomic analyses found that social isolation-related brain changes were spatially correlated with genes associated with mitochondrial function and oxidative stress, and with AD-related down-regulated genes. Taken together, evidence from behavior, neuroimaging and transcriptome revealed that social isolation was an independent risk factor of cognitive decline and dementia. In the context of COVID-19 pandemic which critically exacerbated social isolation, our findings may provide insights into effective intervention strategies especially for old people.

To our knowledge, this is the first large-scale study clarifying the contribution of established risk factors to associations of social isolation and loneliness with dementia. Consistent with previous longitudinal studies,^36, 37^ we found social isolation was associated with 1.28-fold increased risk of developing dementia, which was independent of loneliness and other risk factors.

However, loneliness was not related to incident dementia after full adjustment, owing to 75% of the relationship was attributable to depressive symptoms. Similar results were reported in studies using mortality as the outcome.^23, 38^ Social isolation and loneliness were often weakly correlated,^39^ our results supported that these two might be independent constructs.

We identified multiple brain regions associated with social isolation in the largest sample to date, while no results for loneliness, implying that objective and subjective social relationships possibly have distinct neural bases. In the UK Biobank, socially isolated individuals had the most severe gray matter atrophy in hippocampus, parahippocampus, thalamus, amygdala and temporal cortex, which was not confounded by loneliness and depressive symptoms. The impact of social stressor on hippocampal morphology has been well documented.^40, 41^ Amygdala played an essential role in emotional processing, and was relevant to social network size.^9, 10^ Temporal cortex such as fusiform and superior temporal gyrus was important for social perception.^42, 43^ Experimental manipulation of group size in macaques resulted in variation in the volume of mid-superior temporal sulcus and amygdala.^44^

Cognitive annotation of the significant brain map demonstrated that impaired regions related to social isolation were involved in cognitive processes especially memory and learning. Indeed, we found the volume of these regions was positively associated with cognitive performance, and could partly mediated the prospective association between social isolation at baseline and cognitive performance at the follow-up. Past epidemiological studies have revealed that social isolation was associated with cognitive function in later life,^45^ but evidence on the potential mechanism is sparse. Animal studies suggested that isolation affected cognition via altering the excitatory and inhibitory synaptic density in hippocampus,^46^ and social interaction rescued memory deficit by increasing hippocampal neurogenesis.^47^ The brain reserve hypothesis of dementia posits that larger brains have more neural matter that may increase tolerance of pathology and maintain cognitive function.^48^ Our findings implicated that isolation, as a social stressor, might reduce available brain reserve to accelerate cognitive decline, and finally lead to increased risk of dementia.

The gene expression profile of the social isolation-related cortical changes was enriched in biological processes and KEGG pathways closely linked to dementia. It is well established that mitochondrial dysfunction and oxidative damage is critical in aging and neurodegenerative diseases.^49^ In AD, dysfunctional mitochondria released oxidizing free radicals in the brain and caused considerable oxidative stress.^50^ Animal research showed that impairments of mitochondrial function^51^ and oxidative phosphorylation^52^ preceded the development of AD pathology. We found more down-regulated AD-related genes in the list of positively weighted genes with changes in cortical GMVs. And the expression level of down-regulated genes was significantly associated with cortical changes. These results suggested that genes with reduced brain postmortem transcription in AD were underexpressed in cortical regions with higher levels of changes related to social isolation. Notably, the pattern of brain alterations was obtained from population-based samples, indicating that the brain changes associated with social isolation might precede the diagnosis of dementia.

Some limitations should be mentioned. It was not able to discriminate acute and chronic social isolation in the UK Biobank. Evolutionary theory believes that acute isolation will induce adaptive physiological responses such as increased inflammation and hypothalamic-pituitary-adrenal axis activation to motivate individuals to restore social connections.^53^ While chronic isolation perhaps leads to the set-point adaptations due to the long-term failure to maintain social homeostasis.^54^ The temporal dynamics of social isolation may link to different neural activity and health consequences. Akhter-Khan et al.^55^ reported that transient loneliness was resilient to dementia risk. Consequently, future research should consider the trajectory of social isolation and loneliness. Older adults are particularly vulnerable to COVID-19, and therefore subject to greater social restrictions. It is hypothesized that the virus may increase the risk of cognitive decline and dementia by affecting the brain.^56^ However, the corresponding social isolation measures may also impact the risk of atypical aging and dementia in normal people. Although we currently do not have sufficient data to answer this question, continued monitoring is necessary to investigate the relationship between COVID-19 and dementia.

## Data Availability

This study was conducted under the UK Biobank application number of 19542.

## Data Availability

This study was conducted under the UK Biobank application number of 19542.

## Data Availability

This study was conducted under the UK Biobank application number of 19542.

## ACKNOWLEDGMENTS

This study was conducted under the UK Biobank application number of 19542. Dr Feng was supported by National Key R&D Program of China (No. 2019YFA0709502), National Key R&D Program of China (No. 2018YFC1312904), Shanghai Municipal Science and Technology Major Project (No. 2018SHZDZX01), ZJ Lab, Shanghai Center for Brain Science and Brain-Inspired Technology, and the 111 Project (No. B18015). Dr Sahakian receives funding from a Wellcome Trust Collaborative Award (200181/Z/15/Z), and her research is conducted within the NIHR Cambridge Biomedical Research Centre (BRC) (Mental Health Theme and Neurodegeneration Theme) and the NIHR MedTech and in vitro diagnostic Co-operative (MIC). Dr Cheng was supported by grants from the National Natural Sciences Foundation of China (No. 82071997), the Shanghai Rising-Star Program (No. 21QA1408700) and Natural Science Foundation of Shanghai (No. 18ZR1404400). The funders had no role in study design, data collection and analysis, decision to publish or preparation of the manuscript.

## REFERENCES

1. National Academies of Sciences, Engineering, and Medicine. Social isolation and loneliness in older adults: Opportunities for the health care system. Washington, DC: The National Academies Press; 2020. http://doi.org/10.17226/25663.

2. Prince MJ, Wimo A, Guerchet MM, Ali GC, Wu Y-T, Prina M. World Alzheimer Report 2015: The global impact of dementia: an analysis of prevalence, incidence, cost and trends. London, UK: Alzheimer’s Disease International (ADI); 2015.

3. Penninkilampi R, Casey AN, Singh MF, Brodaty H. The association between social engagement, loneliness, and risk of dementia: a systematic review and meta-analysis. J Alzheimers Dis. 2018;66(4):1619–1633. http://doi.org/10.3233/JAD-180439.

4. Yu B, Steptoe A, Chen Y, Jia X. Social isolation, rather than loneliness, is associated with cognitive decline in older adults: the China Health and Retirement Longitudinal Study. Psychol Med. 2020:1–8. http://doi.org/10.1017/S0033291720001026.

5. Holwerda TJ, Deeg DJ, Beekman AT, et al. Feelings of loneliness, but not social isolation, predict dementia onset: results from the Amsterdam Study of the Elderly (AMSTEL). J Neurol Neurosurg Psychiatry. 2014;85(2):135–142. http://doi.org/10.1136/jnnp-2012-302755.

6. Dunbar RI, Shultz S. Evolution in the social brain. Science. 2007;317(5843):1344–1347. http://doi.org/10.1126/science.1145463.

7. Duzel S, Drewelies J, Gerstorf D, et al. Structural brain correlates of loneliness among older adults. Sci Rep. 2019;9(1):13569. http://doi.org/10.1038/s41598-019-49888-2.

8. Kanai R, Bahrami B, Roylance R, Rees G. Online social network size is reflected in human brain structure. P Roy Soc B-Biol Sci. 2012;279(1732):1327–1334. http://doi.org/10.1098/rspb.2011.1959.

9. Bickart KC, Wright CI, Dautoff RJ, Dickerson BC, Barrett LF. Amygdala volume and social network size in humans. Nat Neurosci. 2011;14(2):163–164. http://doi.org/10.1038/nn.2724.

10. Von Der Heide R, Vyas G, Olson IR. The social network-network: size is predicted by brain structure and function in the amygdala and paralimbic regions. Soc Cogn Affect Neurosci. 2014;9(12):1962–1972. http://doi.org/10.1093/scan/nsu009.

11. Tomova L, Wang KL, Thompson T, et al. Acute social isolation evokes midbrain craving responses similar to hunger. Nat Neurosci. 2020;23(12):1597–1605. http://doi.org/10.1038/s41593-020-00742-z.

12. Inagaki TK, Muscatell KA, Moieni M, et al. Yearning for connection? Loneliness is associated with increased ventral striatum activity to close others. Soc Cogn Affect Neurosci. 2016;11(7):1096–1101. http://doi.org/10.1093/scan/nsv076.

13. Spreng RN, Dimas E, Mwilambwe-Tshilobo L, et al. The default network of the human brain is associated with perceived social isolation. Nat Commun. 2020;11(1):6393. http://doi.org/10.1038/s41467-020-20039-w.

14. Zovetti N, Rossetti MG, Perlini C, Brambilla P, Bellani M. Neuroimaging studies exploring the neural basis of social isolation. Epidemiol Psychiatr Sci. 2021;30:e29. http://doi.org/10.1017/S2045796021000135.

15. Cole SW. Social regulation of human gene expression: mechanisms and implications for public health. Am J Public Health. 2013;103(S1):S84–S92. http://doi.org/10.2105/AJPH.2012.301183.

16. Canli T, Yu L, Yu X, et al. Loneliness 5 years ante-mortem is associated with disease-related differential gene expression in postmortem dorsolateral prefrontal cortex. Transl Psychiatry. 2018;8(1):2. http://doi.org/10.1038/s41398-017-0086-2.

17. Canli T, Wen R, Wang X, et al. Differential transcriptome expression in human nucleus accumbens as a function of loneliness. Mol Psychiatry. 2017;22(7):1069–1078. http://doi.org/10.1038/mp.2016.186.

18. Fornito A, Arnatkeviciute A, Fulcher BD. Bridging the Gap between Connectome and Transcriptome. Trends Cogn Sci. 2019;23(1):34–50. http://doi.org/10.1016/j.tics.2018.10.005.

19. Li J, Seidlitz J, Suckling J, et al. Cortical structural differences in major depressive disorder correlate with cell type-specific transcriptional signatures. Nat Commun. 2021;12(1):1647. http://doi.org/10.1038/s41467-021-21943-5.

20. Zarkali A, McColgan P, Leyland LA, Lees AJ, Rees G, Weil RS. Organisational and neuromodulatory underpinnings of structural-functional connectivity decoupling in patients with Parkinson’s disease. Commun Biol. 2021;4(1):86. http://doi.org/10.1038/s42003-020-01622-9.

21. Hawrylycz MJ, Lein ES, Guillozet-Bongaarts AL, et al. An anatomically comprehensive atlas of the adult human brain transcriptome. Nature. 2012;489(7416):391–399. http://doi.org/10.1038/nature11405.

22. Sudlow C, Gallacher J, Allen N, et al. UK biobank: an open access resource for identifying the causes of a wide range of complex diseases of middle and old age. PLoS Med. 2015;12(3):e1001779. http://doi.org/10.1371/journal.pmed.1001779.

23. Elovainio M, Hakulinen C, Pulkki-Raback L, et al. Contribution of risk factors to excess mortality in isolated and lonely individuals: an analysis of data from the UK Biobank cohort study. Lancet Public heal. 2017;2(6):e260–e266. http://doi.org/10.1016/S2468-2667(17)30075-0.

24. Miller KL, Alfaro-Almagro F, Bangerter NK, et al. Multimodal population brain imaging in the UK Biobank prospective epidemiological study. Nat Neurosci. 2016;19(11):1523–1536. http://doi.org/10.1038/nn.4393.

25. Alfaro-Almagro F, Jenkinson M, Bangerter NK, et al. Image processing and quality control for the first 10,000 brain imaging datasets from UK Biobank. Neuroimage. 2018;166:400–424. http://doi.org/10.1016/j.neuroimage.2017.10.034.

26. Rolls ET, Huang CC, Lin CP, Feng J, Joliot M. Automated anatomical labelling atlas 3. Neuroimage. 2020;206:116189. http://doi.org/10.1016/j.neuroimage.2019.116189.

27. Shen C, Luo Q, Chamberlain SR, et al. What is the link between attention-deficit/hyperactivity disorder and sleep disturbance? A multimodal examination of longitudinal relationships and brain structure using large-scale population-based cohorts. Biol Psychiatry. 2020;88(6):459–469. http://doi.org/10.1016/j.biopsych.2020.03.010.

28. Lin DY, Fleming TR, De Gruttola V. Estimating the proportion of treatment effect explained by a surrogate marker. Stat Med. 1997;16(13):1515–27. http://doi.org/10.1002/(sici)1097-0258(19970715)16:13<1515::aid-sim572>3.0.co;2-1.

29. Therneau TM, Lumley T. Package ‘survival’. R Top Doc. 2015;128(10):28–33.

30. Hayasaka S, Nichols TE. Validating cluster size inference: random field and permutation methods. Neuroimage. 2003;20(4):2343–2356. http://doi.org/10.1016/j.neuroimage.2003.08.003.

31. Tal Yarkoni Rap, Thomas E Nichols, David C Van Essen, Tor D Wager. Large-scale automated synthesis of human fuctional neuroimaging data. Nature Methods. 2011;8(8):665–670. http://doi.org/10.1038/NMETH.1635.

32. Wager TD, Davidson ML, Hughes BL, Lindquist MA, Ochsner KN. Prefrontal-subcortical pathways mediating successful emotion regulation. Neuron. 2008;59(6):1037–1050. http://doi.org/10.1016/j.neuron.2008.09.006.

33. Moreau CA, Urchs SGW, Kuldeep K, et al. Mutations associated with neuropsychiatric conditions delineate functional brain connectivity dimensions contributing to autism and schizophrenia. Nat Commun. 2020;11(1):5272. http://doi.org/10.1038/s41467-020-18997-2.

34. Morgan SE, Seidlitz J, Whitaker KJ, et al. Cortical patterning of abnormal morphometric similarity in psychosis is associated with brain expression of schizophrenia-related genes. Proc Natl Acad Sci U S A. 2019;116(19):9604–9609. http://doi.org/10.1073/pnas.1820754116.

35. Xu M, Zhang DF, Luo RC, et al. A systematic integrated analysis of brain expression profiles reveals YAP1 and other prioritized hub genes as important upstream regulators in Alzheimer’s disease. Alzheimers Dement. 2018;14(2):215–229. http://doi.org/10.1016/j.jalz.2017.08.012.

36. Fratiglioni L, Wang HX, Ericsson K, Maytan M, Winblad B. Influence of social network on occurrence of dementia: a community-based longitudinal study. Lancet. 2000;355(9212):1315–1319. http://doi.org/10.1016/S0140-6736(00)02113-9.

37. Kuiper JS, Zuidersma M, Oude Voshaar RC, et al. Social relationships and risk of dementia: a systematic review and meta-analysis of longitudinal cohort studies. Ageing Res Rev. 2015;22:39–57. http://doi.org/10.1016/j.arr.2015.04.006.

38. Steptoe A, Shankar A, Demakakos P, Wardle J. Social isolation, loneliness, and all-cause mortality in older men and women. Proc Natl Acad Sci U S A. 2013;110(15):5797–801. http://doi.org/10.1073/pnas.1219686110.

39. Coyle CE, Dugan E. Social Isolation, Loneliness and Health Among Older Adults. J Aging Health. 2012;24(8):1346–1363. http://doi.org/10.1177/0898264312460275.

40. Buwalda B, Kole MH, Veenema AH, et al. Long-term effects of social stress on brain and behavior: a focus on hippocampal functioning. Neurosci Biobehav Rev. 2005;29(1):83–97. http://doi.org/10.1016/j.neubiorev.2004.05.005.

41. Stahn AC, Gunga HC, Kohlberg E, Gallinat J, Dinges DF, Kuhn S. Brain Changes in Response to Long Antarctic Expeditions. N Engl J Med. 2019;381(23):2273–2275. http://doi.org/10.1056/NEJMc1904905.

42. Sandi C, Haller J. Stress and the social brain: behavioural effects and neurobiological mechanisms. Nat Rev Neurosci. 2015;16(5):290–304. http://doi.org/10.1038/nrn3918.

43. Allison T, Puce A, McCarthy G. Social perception from visual cues: role of the STS region. Trends Cogn Sci. 2000;4(7):267–278. http://doi.org/10.1016/s1364-6613(00)01501-1.

44. Sallet J, Mars RB, Noonan MP, et al. Social network size affects neural circuits in macaques. Science. 2011;334(6056):697–700. http://doi.org/10.1126/science.1210027.

45. Evans IEM, Martyr A, Collins R, Brayne C, Clare L. Social isolation and cognitive function in later life: a systematic review and meta-analysis. J Alzheimers Dis. 2019;70(S1):S119–S144. http://doi.org/10.3233/JAD-180501.

46. Wang H, Xu X, Xu X, Gao J, Zhang T. Enriched environment and social isolation affect cognition ability via altering excitatory and inhibitory synaptic density in mice hippocampus. Neurochem Res. 2020;45(10):2417–2432. http://doi.org/10.1007/s11064-020-03102-2.

47. Hsiao YH, Hung HC, Chen SH, Gean PW. Social interaction rescues memory deficit in an animal model of Alzheimer’s disease by increasing BDNF-dependent hippocampal neurogenesis. J Neurosci. 2014;34(49):16207–19. http://doi.org/10.1523/JNEUROSCI.0747-14.2014.

48. Stern Y. Cognitive reserve in ageing and Alzheimer’s disease. Lancet Neurol. 2012;11(11):1006–12. http://doi.org/10.1016/S1474-4422(12)70191-6.

49. Beal MF. Mitochondria take center stage in aging and neurodegeneration. Ann Neurol. 2005;58(4):495–505. http://doi.org/10.1002/ana.20624.

50. Querfurth HW, LaFerla FM. Alzheimer’s disease. N Engl J Med. 2010;362(4):329–344. http://doi.org/10.1056/NEJMra0909142.

51. Yao J, Irwin RW, Zhao LQ, Nilsen J, Hamilton RT, Brinton RD. Mitochondrial bioenergetic deficit precedes Alzheimer’s pathology in female mouse model of Alzheimer’s disease. Proc Natl Acad Sci U S A. 2009;106(34):14670–14675. http://doi.org/10.1073/pnas.0903563106.

52. Nunomura A, Perry G, Aliev G, et al. Oxidative damage is the earliest event in Alzheimer disease. J Neuropathol Exp Neurol. 2001;60(8):759–767. http://doi.org/10.1093/jnen/60.8.759.

53. Cacioppo JT, Cacioppo S. Loneliness in the modern age: an evolutionary theory of loneliness (ETL). Adv Exp Soc Psychol. 2018;58:127–197. http://doi.org/10.1016/bs.aesp.2018.03.003.

54. Lee CR, Chen A, Tye KM. The neural circuitry of social homeostasis: Consequences of acute versus chronic social isolation. Cell. 2021;184(10):2794–2795. http://doi.org/10.1016/j.cell.2021.04.044.

55. Akhter-Khan SC, Tao QS, Ang TFA, et al. Associations of loneliness with risk of Alzheimer’s disease dementia in the Framingham Heart Study. Alzheimers Dement. 2021;http://doi.org/10.1002/alz.12327.

56. de Erausquin GA, Snyder H, Carrillo M, et al. The chronic neuropsychiatric sequelae of COVID-19: The need for a prospective study of viral impact on brain functioning. Alzheimers Dement. 2021;http://doi.org/10.1002/alz.12255.

